# DIFFERENT PHENOTYPES OF CHRONIC CONSTIPATION IN MALES AND FEMALES

**DOI:** 10.1101/2024.07.01.24309778

**Authors:** Jerry D. Gardner, George Triadafilopoulos

**Affiliations:** Science for Organizations, 75 DeSilva Island Drive, Mill Valley, CA 94941; Division of Gastroenterology and Hepatology, Stanford University School of Medicine, 420 Broadway Street, Pavilion D, 2nd Floor, Redwood City, CA 94063; Silicon Valley Neuro-gastroenterology and Motility Center, 2490 Hospital Drive, Suite 211, Mountain View, CA, 94040

**Keywords:** Constipation, wireless motility capsule, high-resolution anorectal manometry, pelvic floor dyssynergia, defecation disorder, slow transit constipation, colon transit

## Abstract

**INTRODUCTION:** Patients with chronic constipation exhibit symptoms and motility abnormalities that occur in combinations, but the nature of these combinations has not been characterized.

**METHODS:** We calculated prevalences of combinations of symptoms (abdominal pain, infrequent defecation, incomplete evacuation, straining), abnormal motility test results (prolonged colonic transit time, low anal basal pressure, low anal squeeze pressure, poor rectal sensation, absent balloon expulsion), or both using data from 75 females and 91 males with chronic constipation. We calculated the “Cluster Factor” as observed prevalence of a combination of symptoms, abnormal test results or both divided by the prevalence of the combination due to chance. We calculated the conditional probabilities of combinations of symptoms, abnormal motility test results or both given the prevalence of other members of the same combination.

**RESULTS:** Combinations of symptoms alone or abnormal motility test results alone in both males and females, and for combinations of symptoms plus abnormal motility test results in females, failed to cluster together beyond that attributable to chance alone. Males, however, showed significant clustering. Significant conditional probabilities with symptoms, and with symptoms plus abnormal motility test results was higher in males than females. Significant conditional probabilities with abnormal motility test results were not different between males and females.

**CONCLUSIONS:** Gender-related differences in prevalences of combinations of symptoms and abnormal motility test results, of significant Cluster Factors, and of conditional probabilities indicate that chronic constipation in males reflects a fundamentally different disorder from that in females.

## INTRODUCTION

Chronic constipation is typically assessed by clinical history plus measurements of anorectal motility^1^. Others^2^ measuring symptom severity and mean or median values from motility tests in subjects with chronic constipation observed several instances of significant differences between males and females indicating potentially important gender-related clinical differences in chronic constipation. On the other hand, a study of different tests of evacuatory function in constipated subjects compared combinations of prevalences of abnormal tests to chance combination using the kappa statistic for concordance and reported considerable disagreement among the tests^3^.

Recently, we reported age- and gender-related differences in constipation-related symptoms, colon transit time using a wireless motility capsule, high-resolution anorectal motility, rectal sensation testing, and rectal balloon expulsion testing in patients with chronic constipation^4^. In reviewing the data in that report, we noticed that most subjects had multiple symptoms or multiple abnormal motility test results. For the present analyses, we revisited these cohort data to examine possible relationships among combinations of symptoms, and abnormal motility test results^4,5^. We were particularly interested in comparing observed combinations to possible chance combinations as well as which elements of a combination might be able to predict the occurrence of other elements of the same combination in subjects stratified by gender.

Writing about disorders of brain-gut interaction, Drossman comments that a syndrome is based on symptoms that cluster together^6^. Whereas the diagnosis of functional constipation is based on symptoms^6^, the diagnosis of functional defecation disorders is based on symptoms plus abnormal motility test results^1,6^. In the present analyses, we conducted an unbiased examination of possible clustering of symptoms, abnormal motility test results, or both. Patient-reported symptoms have also been claimed to be poor indicators of underlying pathophysiology^7–9^, and we therefore examined the possibility that elements of a combination of symptoms, abnormal motility test results, or both could provide useful information regarding the occurrence of other elements of these combinations by calculating conditional probabilities.

Our examinations were unbiased in the sense that we examined all possible combinations of 4 different symptoms and 5 different abnormal motility test results from 166 patients that were stratified only by gender and whose only diagnosis was a self-reported complaint of chronic constipation.

## METHODS

The database for the present analyses was created from de-identified symptoms and results from motility tests that were collected systematically from patients with clinically undifferentiated chronic constipation as part of a retrospective, IRB-approved study (El Camino Hospital IRB, Mountain View, CA^5^)

Results for four symptoms (abdominal pain, infrequent defecation, incomplete evacuation, and straining) and five abnormal motility test results (prolonged colonic transit time, CTT, low anal basal pressure, low anal squeeze pressure, poor rectal sensation, and absent balloon expulsion) were recorded for each subject. We stratified subjects based on gender – 91 males: 75 females.

We calculated the prevalence of various combinations of symptoms and abnormal motility test results and presented values from these calculations for a combination of symptoms listed in a row at the top of a table plus a combination of abnormal motility test results listed in a column at the edge of the table.

It is important to note that the fraction of subjects with a particular combination of symptoms or abnormal motility test results can never be numerically greater than the lowest fraction of an element of the combination.

According to probability theory^10^, if the elements of a combination are independent, i.e., due to chance, their combined probability is the product of their individual probabilities. That is, the probability of a combination of independent variables P(A&B) equals P(A) x P(B). If the combination of these variables is greater than chance, the ratio of the observed probability to the chance probability will be greater than 1.0. That is, P(A&B) / (P(A) x P(B)) is greater than 1.0. We examined the possibility that combinations of symptoms, abnormal motility test results or both occurred in clusters by comparing the observed prevalence of a combination to the chance prevalence of the elements of the combination. We calculated the Cluster Factor as the observed prevalence of a combination divided by the chance prevalence of the combination. We defined the Cluster Factor as being statistically significant if the 95% confidence interval for the prevalence of the observed combination did not include the value for the chance prevalence of the elements of the combination.

Also, according to probability theory^10^, the conditional probability of one member of a combination occurring given that the other member of the combination has occurred is calculated by dividing the observed prevalence of the combination by the prevalence of the other member. That is, the conditional probability of A given the probability of B (represented as P(A|B)) is calculated as P(A|B) = P(A&B)/ P(B). Similarly, the conditional probability of B given the probability A (represented as (P(B|A)) is calculated as P(B|A) = P(A&B)/ P(A).

For the present analyses, we calculated the conditional probabilities of combinations of symptoms, abnormal motility test results or both given the prevalence of other members of the same combination. We defined the conditional probability of a variable as being statistically significant if the value for the conditional probability was not included in the bounds of the 95% confidence interval for the prevalence of the variable alone.

Because the present analyses were exploratory, we did not adjust confidence intervals for multiple comparisons.

## RESULTS

Table 1 gives the abbreviations used for the present analyses.

**TABLE 1.** SYMBOLS FOR SYMPTOMS AND ABNORMAL MOTILITY TEST RESULTS.

| TABLE 1. SYMBOLS FOR SYMPTOMS AND ABNORMAL MOTILITY TEST RESULTS. |  |
| --- | --- |
| SYMPTOMS | MOTILITY TEST RESULTS |
| A abdominal pain | J prolonged colonic transit time (CTT) |
| B infrequent defecation | K low anal basal pressure |
| C incomplete evacuation | L low anal squeeze pressure |
| D straining | M poor rectal sensation |
|  | N absent balloon expulsion |

Figure 1 displays the values for prevalence of combinations of symptoms, abnormal motility test results or both in males and females. For both genders, the heat maps indicate that values for prevalence tend to decrease with increasing size of the combinations. Pairwise comparisons of prevalence values indicated that 279 of 511 possible comparisons were higher in males than females (Binomial probability = 0.0041).

**FIGURE 1.**
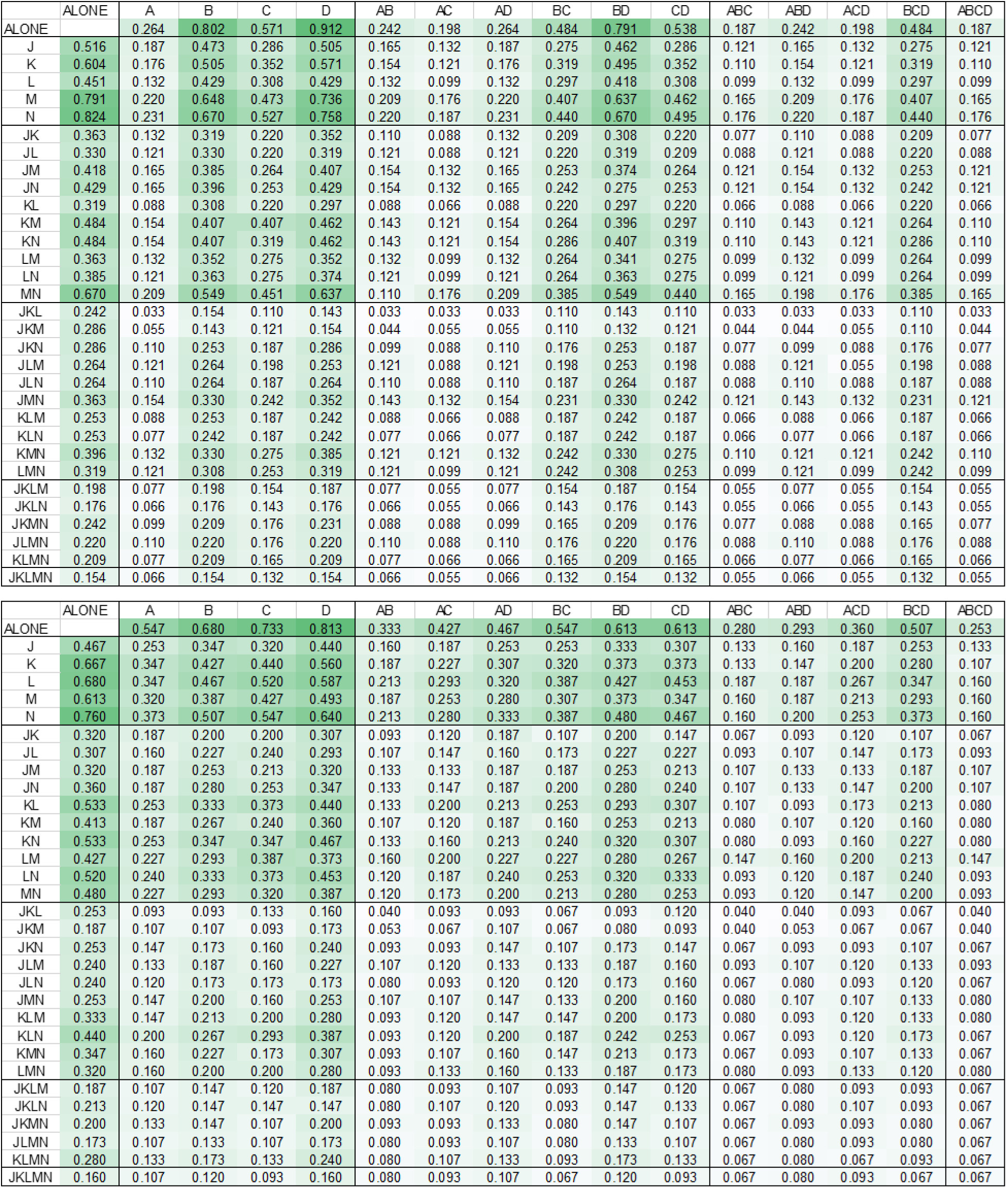
PREVALENCE OF SYMPTOMS, ABNORMAL MOTILITY TEST RESULTS OR BOTH IN SUBJECTS WITH CHRONIC CONSTIPATION. Both panels give values for symptoms and abnormal motility test results from 91 males (top panel) and 75 females (bottom panel). Letters in the top row (A, B, C, D) and left column (J, K, L, M, N) refer to the symptoms and abnormal motility test results given in Table 1. Values in the row and column labeled “ALONE” give individual prevalences for these measures. All other cells give the value for the prevalence of the combination of symptoms in the top row plus abnormal motility test results in the left column. The intensity of the color in the heat map increases with increasing values.

Figure 2 displays the Cluster Factors for the prevalence values in Figure 1. For the 11 possible combinations of symptoms, the Cluster Factors were greater than 1.0 for 11 combinations in males and 9 combinations in females. None of the Cluster Factors for symptoms were statistically significant in males or females because the 95% confidence interval for the prevalence of the observed combination included the value for the chance prevalence of the elements of the combination.

**FIGURE 2.**
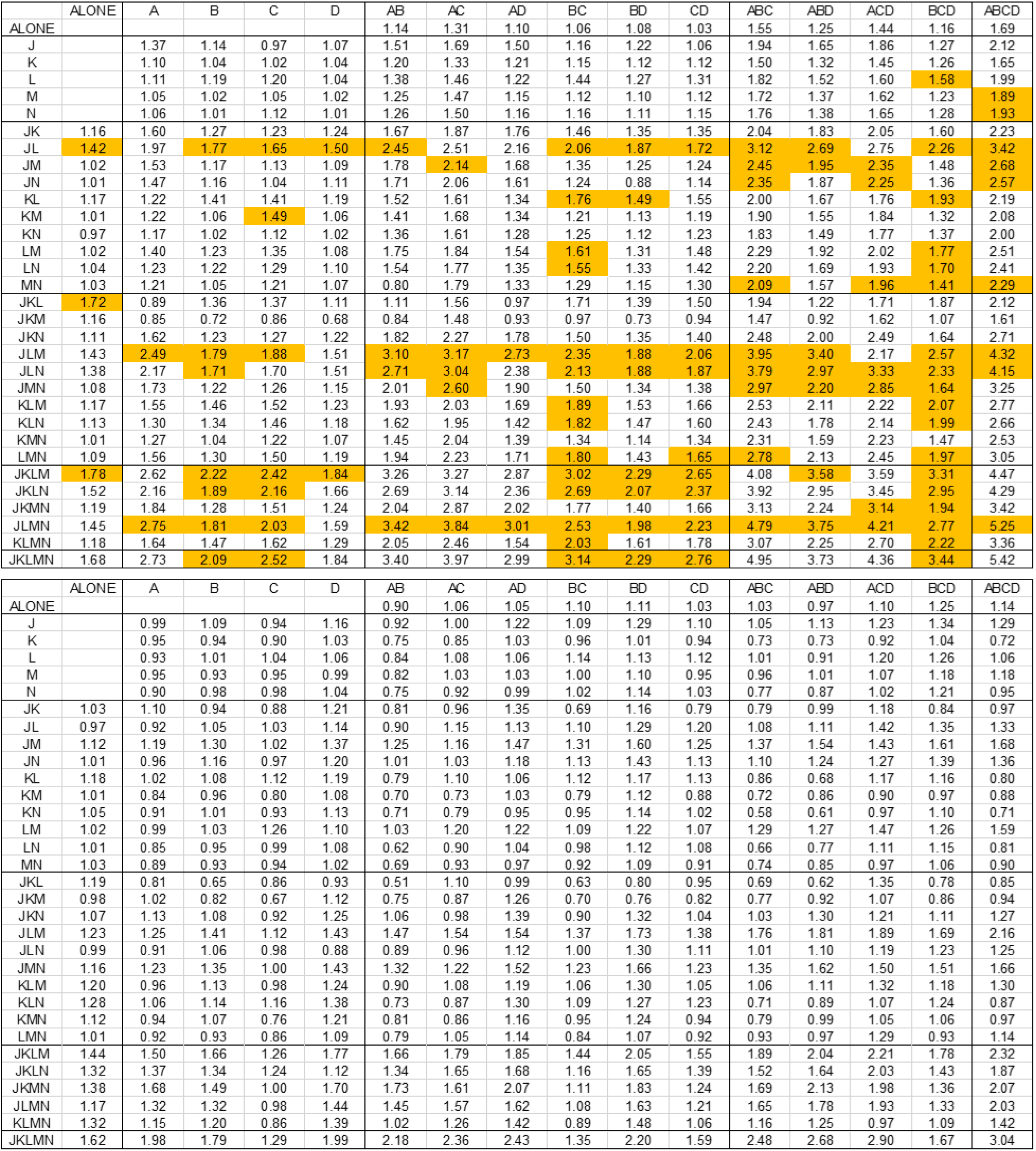
CLUSTER FACTORS FOR THE PREVALENCE OF SYMPTOMS, ABNORMAL MOTILITY TEST RESULTS OR BOTH IN SUBJECTS WITH CHRONIC CONSTIPATION. Both panels give values for symptoms and abnormal motility test results from 91 males (top panel) and 75 females (bottom panel). Letters in the top row (A, B, C, D) and left column (J, K, L, M, N) refer to the symptoms and abnormal motility test results given in Table 1. Values in the row and column labeled “ALONE” give individual cluster factors for prevalences of these measures. All other cells give the value for the cluster factors of the prevalence of the combination of symptoms in the top row plus abnormal motility test results in the left column. The highlighted values indicate cluster factors where the 95% confidence interval of the prevalence did not include the value for the product of the elements of the combination.

For the 26 possible combinations of abnormal motility test results, the Cluster Factors were greater than 1.0 for 25 combinations in males and 23 combinations in females. Three of the Cluster Factors for abnormal motility test results were statistically significant in males, but none in females. The difference between males and females with respect to the number of significant Cluster Factors was not significant (P=0.235 by Fisher’s Exact Test).

For the 465 possible combinations of symptoms plus abnormal motility test results, the Cluster Factors were greater than 1.0 for 450 combinations in males and 306 combinations in females. For males, 112 of the Cluster Factors for combinations of symptoms plus abnormal motility test results were statistically significant, but for females none were statistically significant. There was a significant difference between males and females with respect to the number of Cluster Factors greater than 1.0 as well as the number of significant Cluster Factors (P<0.0001 by Fisher’s Exact Test for both measures).

Figure 3 shows that of the 50 possible conditional probabilities in males, all were higher than the corresponding prevalence alone and 27 were significant in that they were higher than the upper bound of the 95% confidence interval for the corresponding prevalence alone. Of the 50 possible conditional probabilities in females, 39 were higher than the corresponding prevalence alone and 7 were significant. The number of conditional probabilities that were higher than the corresponding prevalence as well as number of significant conditional probabilities were significantly higher in males than females (P=0.0005 and P <0.0001, respectively by Fisher’s Exact Test).

**FIGURE 3.**
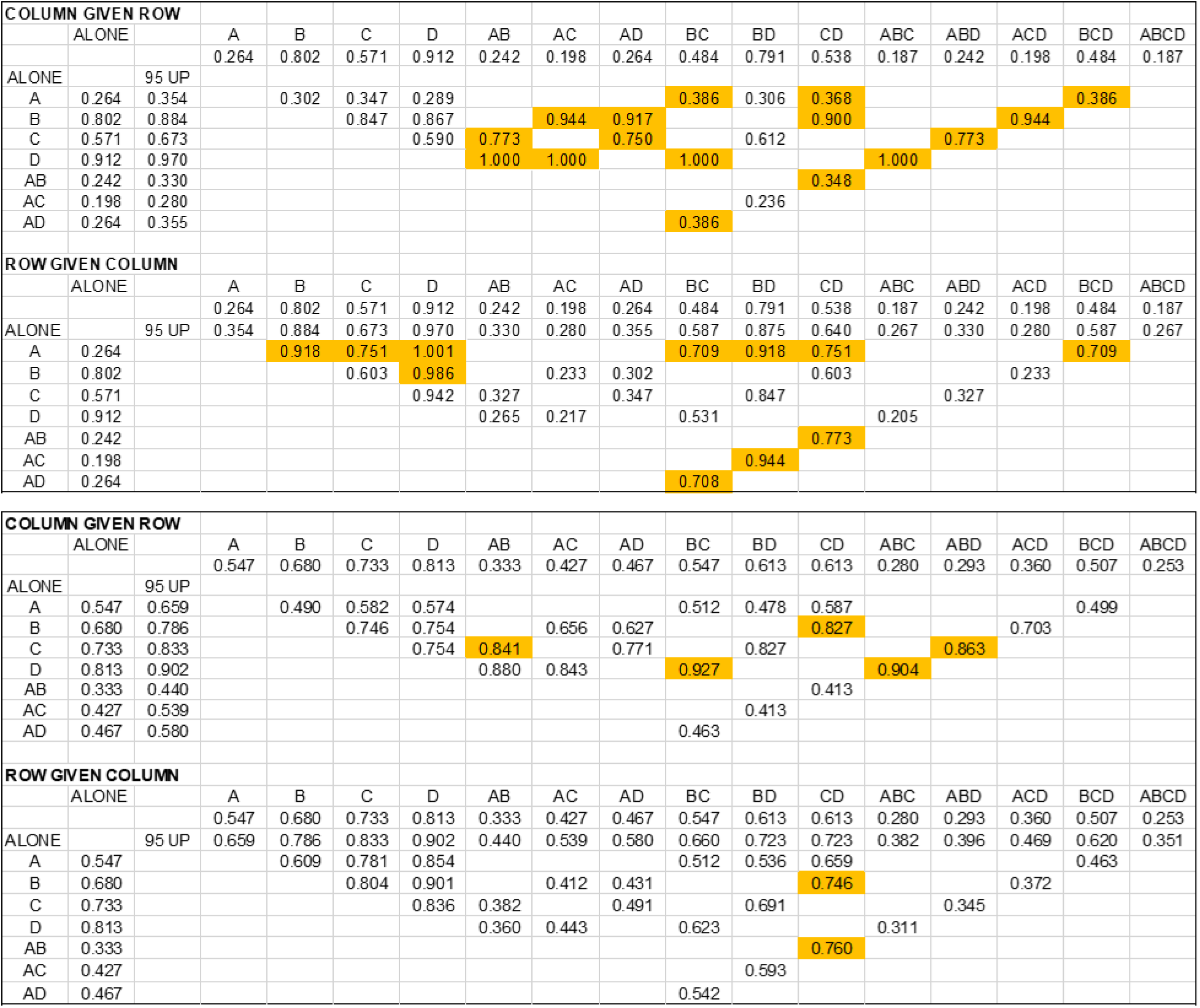
CONDITIONAL PROBABILITIES OF SYMPTOMS PREDICTING OTHER SYMPTOMS IN SUBJECTS WITH CHRONIC CONSTIPATION. Values in the top two panels are from 91 males and in the bottom two panels are from 75 females. Values given are conditional probabilities for each pair of symptoms in each combination calculated using the values for symptoms in the column or row labeled “ALONE” plus the observed prevalence of the corresponding combination given in Figure 1. Letters (A, B, C, D) refer to symptoms given in Table 1. COLUMN GIVEN ROW gives the probability of symptoms in the left column given symptoms in the top row. ROW GIVEN COLUMN gives the probability of symptoms in the top row given symptoms in the left column. Shaded values are higher than the upper bound of the 95% confidence interval of the value for the symptoms given in the column or row labeled “95 UP”.

Figure 4 displays results from males and females with chronic constipation for the conditional probability of one or more abnormal motility test results in a combination given other abnormal motility test results in the combination. Of the 180 possible conditional probabilities in males, 152 were higher than the corresponding prevalence alone and 55 were significant in that they were higher than the upper bound of the 95% confidence interval for the corresponding prevalence alone. Of the 180 possible conditional probabilities in females, 162 were higher than the corresponding prevalence alone and 50 were significant. The number of conditional probabilities that were higher than the corresponding prevalence alone as well as number of significant conditional probabilities were not significantly different between males and females (P=0.1548 and P=0.6249, respectively by Fisher’s Exact Test).

**FIGURE 4.**
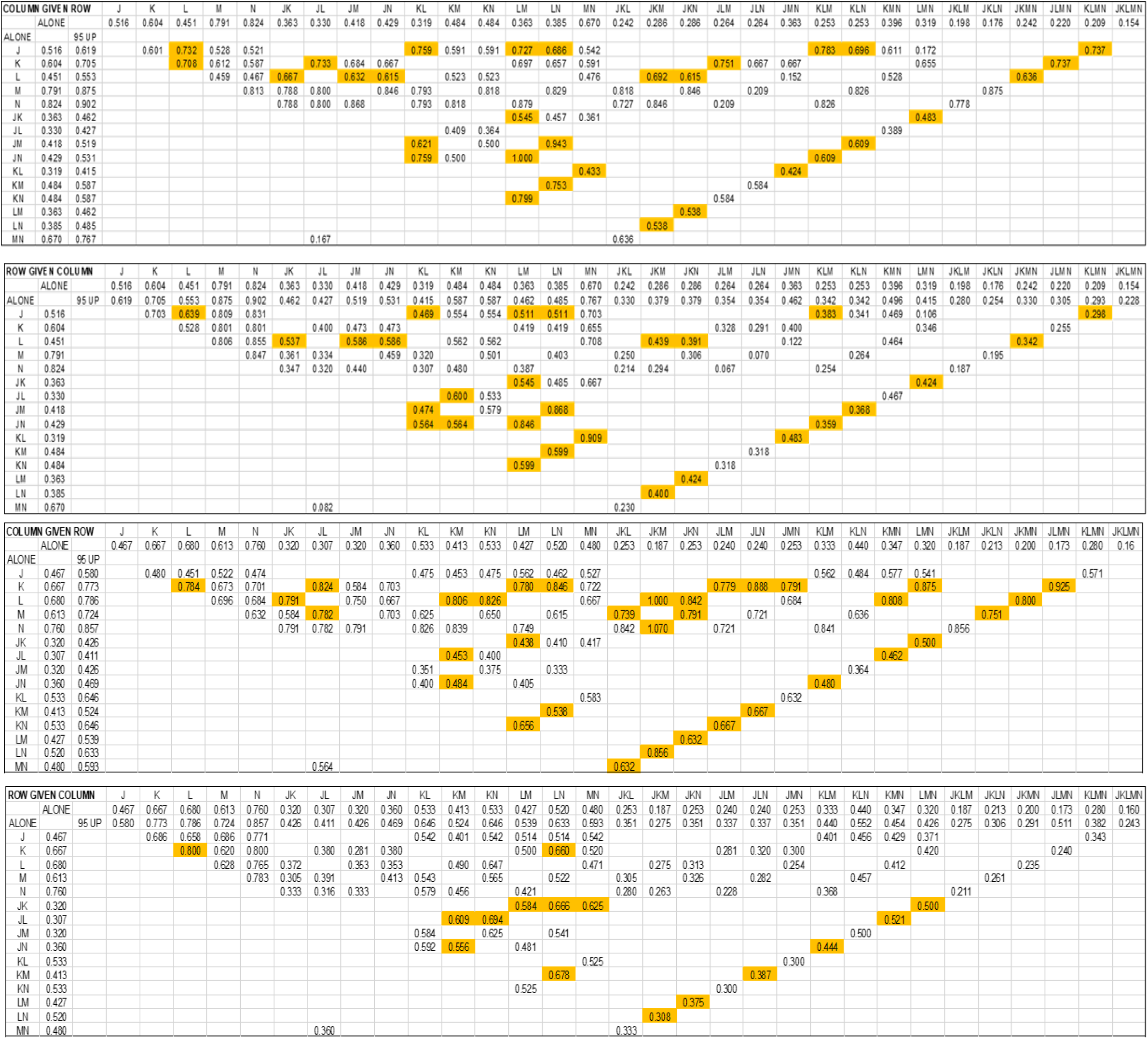
CONDITIONAL PROBABILITIES OF ABNORMAL MOTILITY TEST RESULTS PREDICTING OTHER ABNORMAL TEST RESULTS IN SUBJECTS WITH CHRONIC CONSTIPATION. Values in the top two panels are from 91 males and in the bottom two panels are from 75 females. Values given are conditional probabilities for each pair of abnormal motility test results in a given combination calculated using the values for motility test results in the column or row labeled “ALONE” plus the observed prevalence of the corresponding combination given in Figure 1. Letters (J, K, L, M, N) refer to abnormal motility test results given in Table 1. COLUMN GIVEN ROW gives the probability of test results in the left column given test results in the top row. ROW GIVEN COLUMN gives the probability of test results in the top row given test results in the left column. Shaded values are higher than the upper bound of the 95% confidence interval of the value for the test results given in the column or row labeled “95 UP”.

Figures 5 and 6 display results from males and females with chronic constipation for the conditional probability of one or more abnormal motility test results in a combination given one or more symptoms or vice versa. With 4 different symptoms and 5 different motility tests, there are 465 possible combinations of abnormal motility test results plus symptoms as well as 465 possible conditional probabilities for elements of the combinations.

**FIGURE 5.**
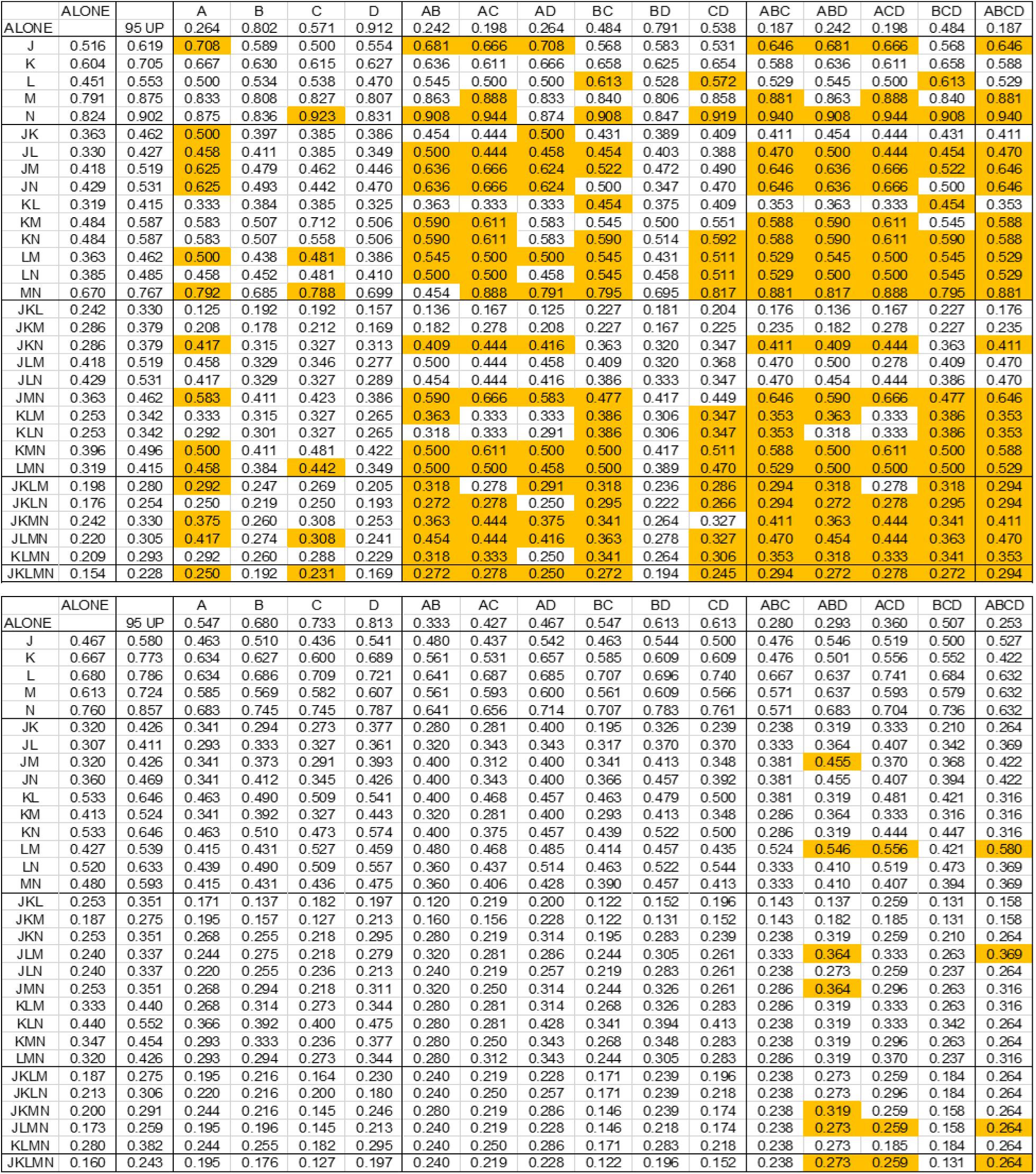
CONDITIONAL PROBABILITIES OF SYMPTOMS PREDICTING ABNORMAL TEST RESULTS IN SUBJECTS WITH CHRONIC CONSTIPATION. Letters in the top row (A, B, C, D) and left column (J, K, L, M, N) refer to the symptoms and abnormal motility test results given in Table 1. Conditional probabilities for each pair of symptoms plus abnormal motility test results in a given combination were calculated using values for symptoms in the row and values for abnormal motility test results in the column labeled “ALONE” plus the observed prevalence of the corresponding combination given in the appropriate panel in Figure 1. Values given are for the probability of abnormal motility test results in the left column given symptoms in the top row. Shaded values are higher than the upper bound of the 95% confidence interval of the value for test results given in the column labeled “95 UP”. Top panel gives results from 91 males and Bottom panel gives results from 75 females.

**FIGURE 6.**
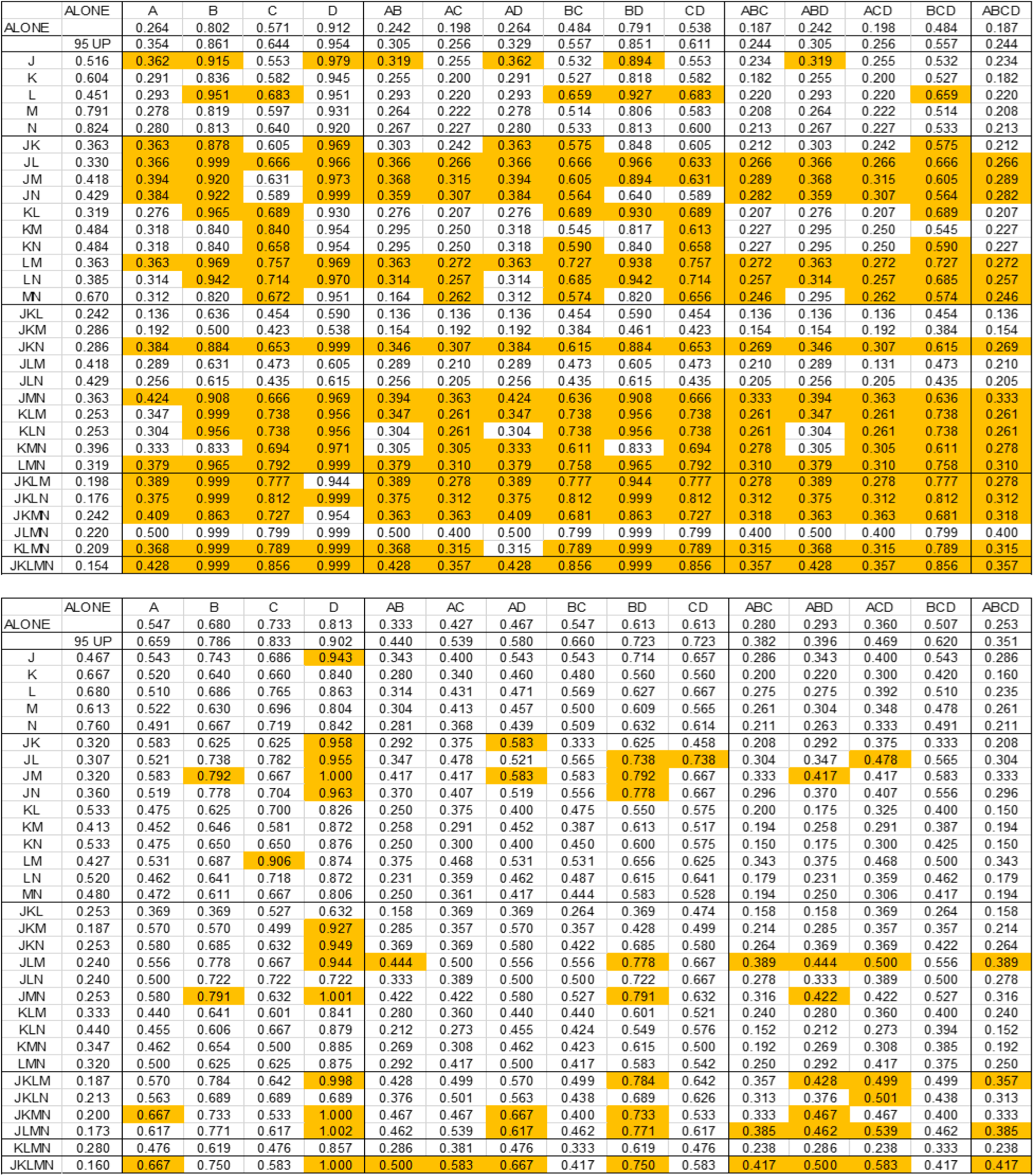
CONDITIONAL PROBABILITIES OF ABNORMAL MOTILITY TEST RESULTS PREDICTING SYMPTOMS IN SUBJECTS WITH CHRONIC CONSTIPATION. Letters in the top row (A, B, C, D) and left column (J, K, L, M, N) refer to the symptoms and abnormal motility test results given in Table 1. Conditional probabilities for each pair of symptoms plus abnormal motility test results in a given combination calculated using values for symptoms in the row and values for abnormal motility test results in the column labeled “ALONE” plus the observed prevalence of the corresponding combination given in the appropriate panel in Figure 1. Values given are for the probability of symptoms in the top row given abnormal motility test results in the left column. Shaded values are higher than the upper bound of the 95% confidence interval of the value for symptoms given in the row labeled “95 UP”. Top panel gives results from 91 males and Bottom panel gives results from 75 females

Figure 5 shows that in males, of the 465 possible conditional probabilities for abnormal test results given symptoms, 413 were higher than the corresponding prevalence alone and 219 were significant in that they were higher than the upper bound of the 95% confidence interval for the corresponding prevalence alone. In females, of the 465 possible conditional probabilities for abnormal test results given symptoms, 205 were higher than the corresponding prevalence alone but only 14 were significant in that they were higher than the upper bound of the 95% confidence interval for the corresponding prevalence alone.

Figure 6 shows that in males, of the 465 possible conditional probabilities for symptoms given abnormal test results, 413 were higher than the corresponding prevalence alone and 277 were significant in that they were higher than the upper bound of the 95% confidence interval for the corresponding prevalence alone. In females, of the 465 possible conditional probabilities for symptoms given abnormal test results, 205 were higher than the corresponding prevalence alone and 59 were significant in that they were higher than the upper bound of the 95% confidence interval for the corresponding prevalence alone.

In both Figure 5 and 6, the number of conditional probabilities that were higher than the corresponding prevalence as well as number of significant conditional probabilities were significantly higher in males than females (both P <0.0001, respectively by Fisher’s Exact Test).

Figures 5 shows that the number of significant abnormal motility test results associated with symptoms tends to increase with the number of symptoms in a combination. For example, in males, symptoms B (infrequent defecation), D (straining) and BD were associated with no significant abnormal motility test results. Symptoms A (abdominal pain) and C (incomplete evacuation) were each associated with 15 and 7 significant abnormal motility test results out of a possible 31 results, respectively. In contrast, combinations of 3 or 4 symptoms were each associated with 20 to 23 significant abnormal motility test results out of a possible 31 results. In females, the only associations of symptoms with significant abnormal motility test results occurred with combinations of symptoms ABD, ACD and ABCD, and the highest number of abnormal motility test results was 7 out of a possible 31 results.

Figures 6 shows that the number of significant symptoms associated with an abnormal motility test results tended to increase with the number of abnormal motility test results in a combination in both males and females. In males, no significant symptoms were associated with K (low anal basal pressure), M (poor rectal sensation), or N (absent balloon expulsion). Abnormal motility test results J (prolonged CTT) and L (low anal squeeze pressure) were associated were each associated with 7 and 6 significant symptoms out of a possible 15 symptoms, respectively. Also, no significant symptoms were associated with abnormal motility test results JL, JKM, JLM, JLN, and JLMN. Increasing numbers of the remaining abnormal motility test results were generally associated with increasing numbers of significant symptoms. Because the number of significant symptoms in females was clearly lower than that in males, females had more combinations of abnormal motility test results that were associated with no significant symptoms and fewer combinations of abnormal motility test results that were associated with significant symptoms.

## DISCUSSION

A characteristic feature of subjects with chronic constipation is their tendency to experience more than one symptom and more than one abnormal motility test result. Some instances may reflect combinations that occur due to chance association. In other instances where the observed prevalence of the combination is greater than that for chance association, the elements of the combination may reflect important features of the pathophysiology of chronic constipation.

In the present analyses, we found that for combinations of symptoms alone or abnormal motility test results alone in both males and females, and for combinations of symptoms plus abnormal motility test results in females, there was no evidence that they cluster together beyond that attributable to chance alone. For males, however, there was clear evidence that combinations of symptoms plus abnormal motility test results show significant clustering indicating that the associations of these variables in males differs in a fundamental way from that in females.

We also examined the possibility that elements of a combination of symptoms, abnormal motility test results, or both could provide useful information regarding the occurrence of other elements of these combinations by calculating conditional probabilities. We found that the conditional probability of elements of a combination was frequently significantly higher than the prevalence of the elements alone indicating that it was possible that elements of a combination could predict the occurrence of other elements of the same combination.

The frequency with which a significant conditional probability occurred with symptoms was significantly higher in males than females. On the other hand, the frequency with this phenomenon occurred with abnormal motility test results was not significantly different between males and females. The frequency of a significant conditional probability of elements of a combination of symptoms plus abnormal motility test results was significantly higher in males than females. These findings indicate that differences between males and females with respect to relationships between symptoms and abnormal motility test results are not attributable to the test results themselves, but instead to the coupling between symptoms and test results. These findings in conjunction with results measuring Cluster Factors offer additional evidence that chronic constipation in females represents a fundamentally different phenotype from that in males

Others^11^ have reported greater variation in males compared to females in biological measures, such as concentrations of various blood constituents that are independent of cultural conditions Our present findings of differences between males and females in the relationships among combinations of symptoms plus abnormal motility test results are consistent with the greater variation in serum laboratory values in males compared to females.

Patient-reported symptoms have been reported to be poor indicators of underlying pathophysiology^7–9^. This conclusion, however, appears to be based on a study of 105 patients, 101 of whom were female^8^, or 60 patients, 56 of whom were female^9^, and our present results show that the poor predictive ability of symptoms was likely attributable to the predominance of females in the studies

In terms of possible impaired treatment responses in females with chronic constipation, no single abdominal, bowel or rectal symptom was found to predict a response to pelvic floor physical therapy in a clinical trial in which 93% of the subjects with chronic constipation were female^9^.

If constipation in females is fundamentally different from that in males, exploring the basis for this difference might provide important insight into the pathophysiology of constipation. Women have a lower anal squeeze pressure, greater perineal descent, longer pudendal nerve terminal motor latency and a greater muscle fiber density than men, while parity leads only to lower squeeze pressure^12^.

In our analyses, the large number of combinations of symptoms and of abnormal motility test with a significant conditional probability of one combination given the prevalence of the other combination raises the possibility of multiple pathophysiologic phenotypes in our cohort, particularly in males. Possibly, different therapeutic interventions can identify these currently unidentified phenotypes. For example, in males with symptom A (abdominal pain) and the abnormal motility test result J (prolonged CTT), the conditional probability of the abnormal motility test result given the symptom or vice versa was higher than the upper bound of the 95% confidence interval for the prevalence of abdominal pain or prolonged CTT alone. These findings raise the possibility that increasing CTT in males with chronic constipation and abdominal pain might ameliorate the abdominal pain. In females with abdominal pain, however, there was no significant association of abdominal pain with any combination of abnormal motility test results that might suggest a possible beneficial treatment. Perhaps other symptoms and motility tests that were not assessed in the present cohort might identify important pathophysiologic relationships in chronic constipation in females.

There are several limitations to the present analyses. Results are from a single clinical practice and will require others to determine the extent to which other patients are exchangeable with patients in the present analyses. Symptoms were assessed with questionnaires, which minimize missing data but circumscribe the clinical condition by failing to include other symptoms (e.g., bloating, distention, or early satiety), that might be associated with constipation. Differences between males and females with respect to symptom frequency may represent gender-related differences in communicating symptom information instead of differences in the actual prevalence of symptoms.

Finally, we did not include analyses of symptom severity even though these data were available.

In conclusion, gender-related differences in prevalences of combinations of symptoms and abnormal motility test results, of significant Cluster Factors, and of conditional probabilities, indicate that chronic constipation in males reflects a fundamentally different disorder from that in females.

## Data Availability

All data produced in the present study are available upon reasonable request to the authors. All results in the present paper can be calculated from the data in Figure 1.

## ACKNOWLEDGEMENTS

The present analyses have been submitted previously to three different journals. Two journals rejected our paper without external review. The third journal rejected our paper with comments from the editors plus an external reviewer. In SUPPLEMENTAL MATERIAL, we have provided the comments from the journals and our responses to the comments from the editors and reviewer from the third journal.

## SUPPLEMENTAL MATERIAL

2-13-24 Neurogastroenterology and Motility

Dear Dr Gardner,

Your manuscript entitled “CLUSTERING OF SYMPTOMS AND ABNORMAL MOTILITY TEST RESULTS IN PATIENTS WITH CHRONIC CONSTIPATION.” has been evaluated by the Editors, and I am sorry to say is not considered suitable for publication in Neurogastroenterology and Motility. We are experiencing an increasing number of submitted manuscripts and to respect the effort authors put into their manuscripts we do not send out manuscripts for review when they are highly unlikely to be accepted. In this case the editors feel that the paper has low priority

I am sorry for the negative decision and hope it will not deter you from sending future manuscripts to the Journal.

Yours sincerely,

Christopher Black

Associate Editor

Frank Zerbib

Editor

Maura Corsetti

Editor in Chief

Neurogastroenterology and Motility

COMMENT: This version of our manuscript contained only the data for Cluster Factors illustrated in Table1 and Figures 1 and 2 of the current manuscript.

4-4-24 American Journal of Gastroenterology

AJG-24-0708

DIFFERENTIAL PHENOTYPES OF CHRONIC CONSTIPATION IN MALES AND FEMALES

Dr. Gardner,

Your manuscript has been reviewed at the editorial office. Unfortunately, despite its undoubted interest, it is felt that it is unlikely to achieve an adequate priority for publication.

Dear Dr. Gardner,

We found this manuscript novel but think it is better suited for a motility-based journal rather than AJG as the study design limits the interpretation and external generalizability of your findings.

We receive far more manuscripts than our reviewers can handle and must reject some papers without review based on the priority for publication. Decisions regarding priority are made by the Editors and Editorial Board based on the subject matter, the relevance to our readership, study design, originality, substantive editorial concerns, and the submission of other similar studies.

We do appreciate your thoughtfulness in submitting your manuscript to the American Journal of Gastroenterology for consideration.

Yours sincerely,

Baha Moshiree, MD, MSc, FACG

Associate Editor

Jasmohan Bajaj, MD, MS, FACG

Millie Long, MD, MPH, FACG

Co-Editors-in-Chief

The American Journal of Gastroenterology

COMMENT: This version of our manuscript contained the same Table and Figures as the current manuscript.

6-8-24 Clinical Gastroenterology and Hepatology

Ref.: Ms. No. CGH-D-24-01229

Title: “DIFFERENT PHENOTYPES OF CHRONIC CONSTIPATION IN MALES AND FEMALES”

*Clinical Gastroenterology and Hepatology*

Jun 08 2024 05:47AM

Dear Dr. Gardner,

We regret to inform you that your manuscript entitled “DIFFERENT PHENOTYPES OF CHRONIC CONSTIPATION IN MALES AND FEMALES” was not accepted for publication in *Clinical Gastroenterology and Hepatology*. The manuscript was externally reviewed and was also reviewed by the Associate Editor and the Board of Editors. Substantive issues and concerns were raised that--combined with constraints on the number of papers that can be published--precluded assigning a high priority score to your manuscript to merit publication. *CGH* receives over 2,300 submissions a year and we can only accept a small fraction of these papers.

We thank you for having allowed us to consider this work for publication in *Clinical Gastroenterology and Hepatology*. We hope you find these comments useful when submitting your paper to another journal. We look forward to future submissions of other manuscripts from your group.

Sincerely yours,

Anthony Lembo, MD

Associate Editor

Charles J. Kahi, MD, MSc

Editor-in-Chief

*Clinical Gastroenterology and Hepatology*

Editor and Reviewer comments:

Unfortunately we are unable to accept your article in CGH. We hope you find the comments made by the reviewer helpful for your future submission. We also recommend including a table with patient demographics. Ideally, additionally we recommend future analyses with all constipation symptoms rather than the selected symptoms.

REPLY:

We agree that including a Table with demographics would have been helpful in the present manuscript; however, adding this Table would have exceeded the limit the Journal allows for the number of Tables and Figures in a manuscript. We have provided the demographic information for the subjects in our database in reference 4 in our list of references in the current manuscript.

Good scientific practices indicate that when analyses are based on a database, the database must be locked before any analyses are conducted. Once the database is locked, no additions, subtractions or modifications are allowed. We believe that it would not be appropriate to conduct future analyses using our database with other constipation symptoms. On the other hand, in the Discussion of our paper we have suggested symptoms that could be included in future databases containing symptoms from subjects with chronic constipation.

Reviewer #1: Comments to the Editors:

In this manuscript, the authors used cross-sectional data from a single center of patients presenting with chronic constipation to determine if certain phenotypes of patients cluster together based on symptom report and findings on motility testing (colonic transit testing and anorectal manometry). They examined 4 different symptoms and 5 different abnormal motility parameters from 166 total patients. They used probability theory to determine that males—but not females—had significant clustering, which they suggested indicates major differences in constipation by sex.

1. The authors chose 4 symptoms and 5 motility testing parameters for their analysis of clustering but there is little explanation on how these 4 symptoms (of many possible symptoms of constipation) were chosen and how they were collected (i.e. by validated questionnaires).

REPLY:

Symptoms (lower abdominal pain, infrequent defecation of hard stools, sense of incomplete evacuation of stools, and straining during defecation) were collected using a standard questionnaire that asked each patient to report “yes” or “no” for each symptom. A patient who reported a symptom was asked to also rate its severity as “mild”, “moderate” or “severe” (absent = 0, mild = 1, moderate = 2, and severe = 3). This questionnaire has been extensively used and reported previously (Rosa-E-Silva L, Gerson L, Davila M, Triadafilopoulos G. Clinical, radiologic, and manometric characteristics of chronic intestinal dysmotility: the Stanford experience. Clin Gastroenterol Hepatol. 2006; 4: 866-73. (PMID: 16797243).

Similarly, it is not clear how the motility testing was performed. Looking back at their previous publication (citation #5), it would appear that patients underwent wireless motility capsule testing and London protocol anorectal manometry. Although CTT is a reasonable parameter, how were the anorectal manometry parameters chosen?

REPLY:

All parameters of the London protocol anorectal manometry were recorded and specifically, the IAPWG classification (part 3): Disorders of recto-anal coordination, that requires the use of both balloon expulsion test and anorectal manometry.

For example, would not the presence of dyssynergia be relevant since this defines (with balloon expulsion testing) who is referred for biofeedback therapy?

REPLY:

Yes, please see above.

2. When the authors report “absent balloon expulsion”, do they mean >60 seconds or a complete inability to expel the balloon?

REPLY:

In Reference 4, we reported that “absent balloon expulsion” means greater than 60 seconds, not lack of expulsion.

REPLY TO COMMENTS 3-5:

We believe that the Reviewer’s comments 3, 4 and 5 reflect a fundamental misunderstanding of our analyses. Each Reviewer’s comment as well as the references in Comment 4 refer to clusters or groups of <u>patients</u>. In contrast, our analyses examined possible clusters or relationships of <u>symptoms, abnormal motility test results or both</u> in subjects with chronic constipation stratified by gender. Our findings show which combinations of variables occur significantly more frequently than chance associations, and which variables can predict the occurrence of other variables as well as the strength of the prediction. We stratified by gender because as we point out in the Introduction to our paper, we (reference 4) and others (reference 2) have reported gender-related differences in symptoms and motility test results in patients with constipation.

3. Do the authors have information on IBS diagnoses among those presenting with constipation? Although it can be argued that IBS-C and functional constipation lie on a spectrum, other analyses have shown that IBS patients tend to cluster together.

4. Similar attempts to define clusters of DGBI patients have been performed before for both chronic constipation (PMID: 38396355) and IBS (PMID: PMID: 36858142) using latent profile or latent class analyses and other studies have used principal components analyses. I have not seen this methodology used previously. How does this probability theory compare to other methods of determining latent phenotypes and what are its relative advantages?

5. The authors noted that clusters can be determined in male patients but not females. Why the choice to stratify by sex and not other variables that may similarly show the ability to better define these constipation phenotypes?

6. Among only 166 patients, the analysis includes hundreds—if not more—of comparisons. Are there any concerns about false discovery rate in this type of analysis?

REPLY:

We agree with the Reviewer that the false discovery rate is an important issue regarding our analyses. All of our major findings are based on confidence intervals and adjusting confidence intervals for multiple comparisons is more complicated than adjusting P-values for multiple comparisons (see Journal American Statistical Association 2005; 100:71-81. False Discovery Rate–Adjusted Multiple Confidence Intervals for Selected Parameters}. Since our analyses were exploratory, we did not adjust the confidence intervals for multiple comparisons.

7. The authors note that several phenotypes are found in male constipation patients. Can the authors opine on who constitutes these significant phenotypes—especially the very strong associations?

REPLY:

We agree with the Reviewer that the possibility of different phenotypes of male subjects with constipation is important. Unfortunately, with the 4,000-word limitation for papers published in Clinical Gastroenterology and Hepatology we were unable to include results from additional analyses in our paper.

